# Report – Cost and Clinical Utility of WES and WGS in pediatric patients with suspected genetic disease

**DOI:** 10.1101/2022.10.10.22280925

**Authors:** Michael P. Douglas, Patricia A. Deverka, Bruce Gelb, Bart Ferket, Kristen Hassmiller Lich, Hadley Stevens Smith, Mary Norton, Jonathan Berg, Anne Slavotinek, Lucia Hindorff, Kathryn Phillips

## Abstract

Payer coverage for Exome Sequencing (ES) is becoming commonplace (albeit in some cases with prior authorization restrictions), coverage for Genome Sequencing (GS) is rare, with most payers considering it as investigational and not medically necessary. Previous studies had identified several concerns and challenges from the payer perspective. **The objective of this study is to conduct a targeted literature review of the evidence describing the cost and clinical utility of GS and ES compared to standard of care (SoC) testing in children ≤18 years with suspected genetic disease**.

We conducted a systematic literature review to identify evidence for cost, diagnostic utility and clinical utility between SoC, ES, and GS in children (0-18 years) with suspected genetic diseases. We also identified list prices for individual tests from Concert Genetics. Descriptive analyses were conducted for cost and comparative effectiveness data.

We identified five studies on costs of ES and GS, as well as data from concert genetics, and 62 studies of comparative effectiveness of GS or ES in pediatric patients <18 yrs. with suspected genetic disease. We identified 2 reviews/systematic literature review, one cost savings, one micro-costing, and one cost-effectiveness analysis, in addition to the list prices from Concert Genetics which support the range of costs in the articles. List prices ranged from an average of $2264 (small panel) to $7245 for a rapid ES trio. Nearly half the studies (28/63; 44%) only reported diagnostic yield, and 18 studies with NICU/PICU patients and 18 studies of outpatients that reported a change in management. The change in management was very heterogeneous ranging from 24% to 83% depending on the patients’ suspected disorder and test type (e.g., rapid GS, rapid ES, GS). We were unable to determine if any of the GS test types (ultra-rapid GS or rapid GS or GS, range 24% - 83%) provided a greater change in clinical management rate than the ES test types (rapid ES or ES, range 35% - 83%).

There is limited evidence of GS over ES in the ability to effect change in management. Available evidence suggests that rapid over standard GS or ES is of greater benefit in the NICU/PICU setting vs. outpatient setting. Future clinical studies must include both diagnostic yield and clinical management outcomes in order to provide stakeholders with the necessary evidence to support decision-making on implementation and coverage.

## Introduction

While payer coverage for Exome Sequencing (ES) is becoming commonplace (albeit in some cases with prior authorization restrictions), coverage for Genome Sequencing (GS) is rare, with most payers considering it as investigational and not medically necessary. We identified the main concerns and challenges to coverage of GS vs. ES from the payer perspective in two previous studies. [Trosman 2019, Phillips 2020] These studies identified 5 concerns and challenges: 1. lab quality concerns, 2. more variants of unknown significances (VUS) with GS than ES, 3. need for provider education to avoid misuse of ES and GS, 4. costs of GS are higher than ES and both tests cost more than multi-gene panels, and 5. lack of clinical utility data to justify use of GS vs. ES. This report will focus on costs and evidence of clinical utility as these are both necessary for evaluation of test value.

Our report excludes three of the concerns or challenges. First, the <u>concern for lab quality</u> which is primarily addressed in evidence of analytic validity. This concern, while relevant for GS and ES, is also relevant for all Next Generation Sequencing (NGS)-based testing. This concern is directly addressed via the FDA approval process of sequencing machines (e.g., Illumina has approval for their MiSeqDx Sequencer) [Collins, 2013] or through the Clinical Laboratory Improvement Amendments (CLIA) which regulates all laboratory testing performed on humans. [CMS CLIA website - https://www.cms.gov/Regulations-and-Guidance/Legislation/CLIA] Furthermore, any Next Generation Sequencing (NGS)-identified variant of concern should undergo verification using Sanger Sequencing prior to use in clinical decision making. Second, the <u>concern of more VUS with GS than ES</u> is a false assumption in clinical sequencing. While GS generates more sequence data than ES, the VUS outside of the exome have impact in the protein synthesis. VUS are more likely to be identified in non-European Caucasian populations such as African or Asian due to limited or lack of data. Third, the <u>concern for the need of provider education to avoid misuse of GS vs. ES</u>, while valid, also applies to all types of NGS-based testing and laboratory testing. This concern can be addressed by training physicians, the use of prior authorization, and/or the requirement for tests to be ordered by a genetic specialist(s) within the payers’ medical policy.

**The objective of this study is to conduct a targeted literature review of the evidence describing the cost and clinical utility of GS and ES compared to standard of care testing in children ≤18 years with suspected genetic disease**. The concern payers have for the <u>cost of GS vs. ES</u> or other tests (e.g., NGS panels, chromosomal micro-array [CMA]) is valid with GS costing approximately 2x the cost of ES. The main concern with the economic value of GS vs. ES is how this is perceived. Grosse (GIM 2021) describes the varying ways of interpreting cost vs. price and how they differ. Furthermore, they describe how these impact cost-effectiveness analyses, which are also dependent on the clinical context and payer setting. The main concern is <u>evidence of clinical utility of GS and ES</u>. Previous studies, a systematic literature review by Stevens Smith et al (2019), and a meta-analysis by Clark (2018) describe the clinical utility of GS and ES, with both concluding there is limited/no evidence that GS is superior at diagnosis or effecting a change in management or improved clinical outcomes in patients with suspected genetic disorders. In many cases, the definition of clinical utility is different by stakeholder. For example, a patient (or parents of a patient) may find value in the diagnosis of condition even if there is not the ability to change the clinical management or outcome (e.g., morbidity or mortality). A physician may find similar value as they no longer have to attempt to provide a diagnosis to the parents (e.g., ending the diagnostic odyssey), while a payer may not consider this an appropriate endpoint to warrant paying for GS above ES, NGS panels, or CMA. The definition most payers use for clinical utility is the change clinical management that results in a change in health outcome (e.g., decreased morbidity or mortality). In this report, we focus on the comparative effectiveness of clinical utility for diagnostic yield, change in clinical management and change in health outcomes with the use of GS and ES to diagnose children (0-18 years) with suspected genetic disease in both the inpatient (NICU/PICU) and outpatient settings.

## Methods

Our objective was to identify evidence to answer the following research questions:

- What are the costs of ES compared to SoC testing (CMA, panels) in children (0-18 years) with suspected genetic disease?
- What are the costs of GS compared to SoC testing (ES + CMA) in children (0-18 years) with suspected genetic disease?
- What is the diagnostic utility of ES compared to SoC testing (CMA, panels) in children (0-18 years) with suspected genetic disease?
- What is the clinical utility of GS compared to SoC testing (CMA, panels) in children (0-18 years) with suspected genetic disease?
- What is the diagnostic utility of ES compared to GS in children (0-18 years) with suspected genetic disease?
- What is the clinical utility of ES compared to GS in children (0-18 years) with suspected genetic disease?

We conducted a literature review to identify original research articles pertaining to the costs of GS and ES, and comparative effectiveness of the diagnostic yield and clinical utility of GS and ES. We also searched the Concert Genetics website for list prices of ES and GS in the US. We formulated search strategies (MD/PD) and reviewed these with an experienced methodologist (Jennifer Malinowski, ACMG) (Supplemental Table 1: Search Strategies of Cost Studies and Supplemental Table 2: Search Strategies of Comparative Effectiveness Studies).

**TABLE 1:** Cost, and List Price of Rapid & Standard GS and ES (singleton and trio), CMA, and panels.

| Reference<br>(article<br>type) | Rapid GS |  | Rapid ES |  | GS |  | ES |  | CMA | Panels |
| --- | --- | --- | --- | --- | --- | --- | --- | --- | --- | --- |
|  | Single | Trio | Single | Trio | Single | Trio | Single | Trio |  |  |
| Chrystoja<br>2014<br>(Review<br>article) | N/A | N/A | N/A | N/A | \$6500<br>(UT<br>Austin<br>list price) | N/A | N/A | N/A | N/A | N/A |
| Joshi 2016<br>(Cost<br>Savings) | N/A | N/A | N/A | N/A | N/A | N/A | N/A | \$6100 | \$1110 | Epilepsy<br>Panel<br>\$1500 -<br>\$6000<br>(proband<br>only) |
| Sabatini<br>2016<br>(Micro-costing) | N/A | N/A | N/A | N/A | N/A | N/A | \$2428 to<br>\$3388 | N/A | N/A | X-linked<br>Panel<br>\$914;<br>Hearing<br>Loss<br>Panel<br>\$1048 –<br>1949);<br>Clinical<br>Exome<br>Panel<br>(~500<br>genes)<br>\$1499 |
| Schwarze<br>2018<br>(SLR) | N/A | N/A | N/A | N/A | \$1,906 to<br>\$24,810 | N/A | \$555 to<br>\$5,169 | \$3,825 to<br>\$9,304 | N/A | N/A |
| Lavelle<br>2022<br>(CEA) | N/A | \$12,000<br>(\$9,000-<br>\$15,000) | N/A | \$10,320<br>(\$6,720-<br>\$13,920) | N/A | \$10,450<br>(\$7,008-<br>\$14,304) | N/A | \$8,112<br>(\$6,720-<br>\$10,560) | \$2154<sup>2</sup> | N/A |
| **List Price<sup>1</sup>** | N/A | N/A | \$3926.67<br>(\$1490 -<br>\$8000,<br>SD<br>\$3550.22) | \$7245<br>(\$2890 -<br>\$11600,<br>SD<br>\$6158.90) | \$4381<br>(\$2490 -<br>\$7395,<br>SD<br>\$2022.21) | \$6145<br>(\$4990 -<br>\$7300,<br>SD<br>\$1633.42) | \$3113.50<br>(\$1095 -<br>\$5600,<br>SD<br>\$1595.90) | \$4230<br>(\$1500 –<br>\$8000,<br>SD<br>\$2350.35) | \$1565<br>(\$350 -<br>\$2184,<br>SD<br>\$485) | \$2264<br>(\$1595 -<br>\$3000,<br>SD<br>\$529) |

<sup>1</sup> List Prices found on Concert Genetics website were averaged for each type of test. The minimum and maximum are in parentheses with standard deviation.
<sup>2</sup> Genetic testing performed on patients with suspected genetic conditions prior to diagnosis
Abbreviations: N/A = not available

**TABLE 2.** Total Studies by Setting (N = 63 studies, 65 test types)

|  | **Ultra-rapid GS** | **Rapid GS** | **Rapid ES** | **GS** | **ES** |
| --- | --- | --- | --- | --- | --- |
| **Inpatient** | 1 | 10 | 5 | 1 | 1 |
| **Outpatient** |  |  |  | 6 | 41 |

NOTES:
\*Soden is reported twice – once as rapid GS in NICU and once as ES in outpatient
\*\*Meng is reported twice – once as rapid ES in NICU/PICU and once as ES in outpatient

### Inclusion/Exclusion Criteria

- Cost Studies: ES or GS in pediatric setting with results reported in USD.
- Comparative Effectiveness Studies: ES or GS (alone or as a comparison to each other, NGS panel, or SOC) in pediatric setting.
- Limits: Humans, English

### Article Selection

Articles were selected for inclusion in the following manner: One author (MD) screened the titles/abstracts followed by a team of authors that reviewed the full text of the articles (Cost articles – MD/PD; Comparative Effectiveness articles – MN/MD). Additional articles were included using snowball method of reviewing references of included articles.

### Data Abstraction

Teams of authors (MD, PD, MN) abstracted the key variables from included articles. One author (MD) conducted searches and compiled cost data from the Concert Genetics website (www.concertgenetics.com) for GS, ES, NGS Panels, and CMA list prices.

### Data Analysis

Descriptive analyses were conducted for cost and comparative effectiveness data.

## Results

We identified five studies on costs of ES and GS, as well as data from concert genetics, and 62 studies of comparative effectiveness of GS or ES in pediatric patients ≤18 yrs. with suspected genetic disease.

### Cost

We identified 2 reviews/systematic literature review, one cost savings, one micro-costing, and one cost-effectiveness analysis, in addition to the list prices from Concert Genetics which support the range of costs in the articles identified (Table 1). The articles reported costs in a variety of manners:

- list price from one lab (UT Austin) (Chrystoja, 2014)
- prices from three labs (not defined) (Sabatini, 2018)
- price from one lab (Iowa Institute of Human Genetics) (Joshi, 2016)
- costs reported in literature (not defined) (Schwarze, 2018)
- CMS reimbursement rates (Lavelle, 2022)
- List prices from Concert Genetics (this study)

The cost of sequencing is as expected with GS being more expensive than ES which is more expensive than CMA which is similar in price to NGS panels. The cost of GS and ES trios (singleton and 2 parents) is approximately 1.5 times the cost of their respective singleton (proband). The range of prices for a particular test (e.g., GS) varies widely, most likely due to how costs were measured in specific institutions.

### Comparative Effectiveness

We included 63 studies on the comparative effectiveness of GS or ES in pediatric patients with suspected genetic disorders (see Supplemental Figure 1: PRISMA Diagram of Comparative Effectiveness Studies). Some studies included testing conducted in both inpatient and outpatient settings, so the total number of tests studied = 65 (see Table 1). ES in the outpatient setting was the most frequently studied test (41/65), with GS only evaluated as an outpatient test in 6/65. Rapid GS and ES are primarily evaluated in the inpatient (NICU/PICU) setting.

### Diagnostic Yield

Nearly half the studies (28/63; 44%) only reported diagnostic yield and did not address the outcome of the test(s) ability to effect change in management or clinical benefits/harms (See Table 3). All of these studies evaluated outpatient testing and vast majority (24/28) evaluated only ES. There was broad geographic representation in these studies with 43% coming from the US, and others from Canada, The Netherlands, United Kingdom, Saudi Arabia, Qatar, Spain, China, Japan, and Taiwan. The sample size varied between studies with the majority of studies less than 1000 patients with heterogenous comparators (previous genetic and non-genetic testing, CMA, panel) and heterogenous methods for identifying study population. There was a consistent finding that ES proband had higher diagnostic yields than standard of care testing, but depends on the pre-test likelihood of genetic disease, with ES trios having a similar range of diagnostic yields as ES of proband. There were too few studies to conclude if GS has higher diagnostic yield than ES + CMA or other previous genetic and non-genetic testing.

**TABLE 3.**
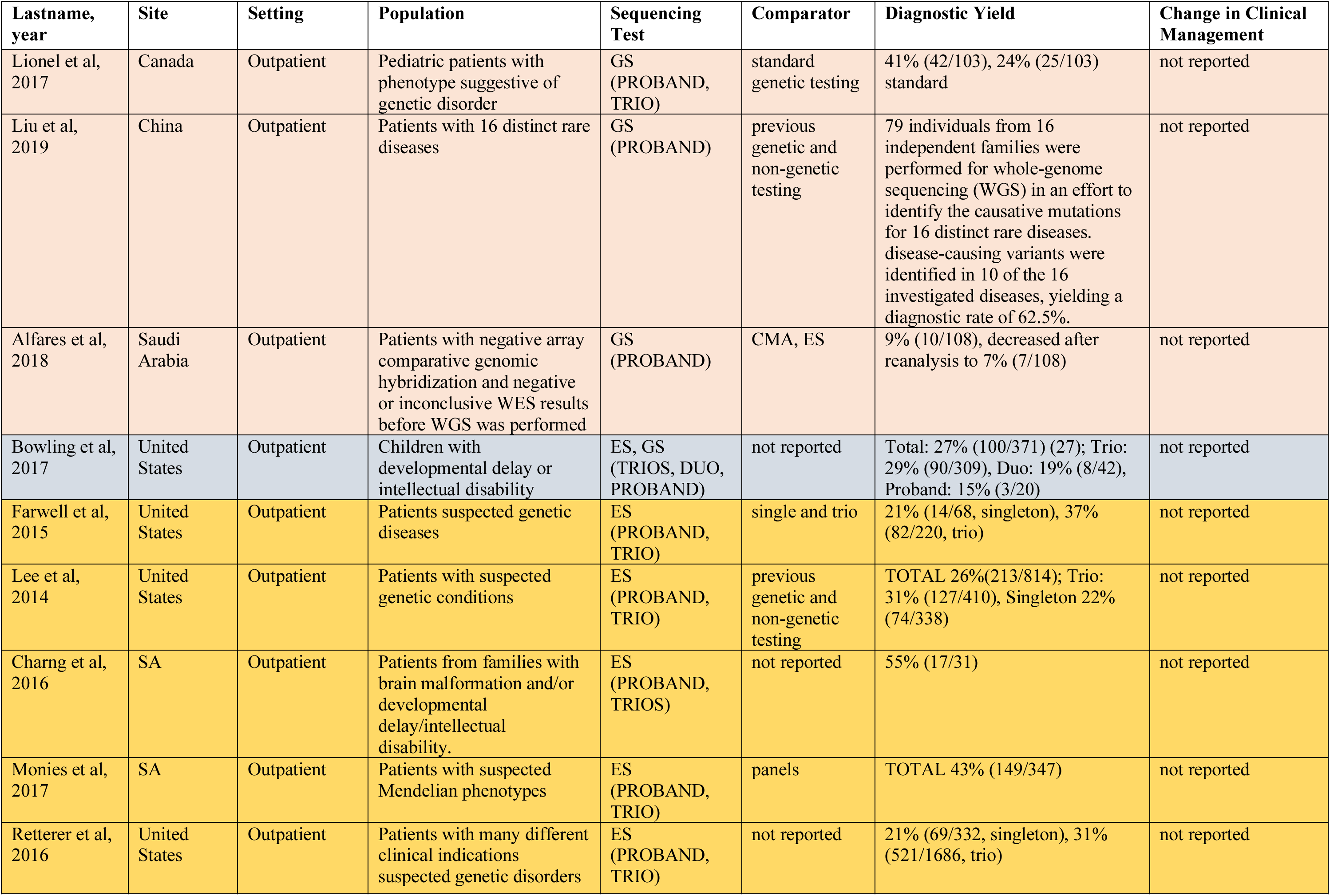
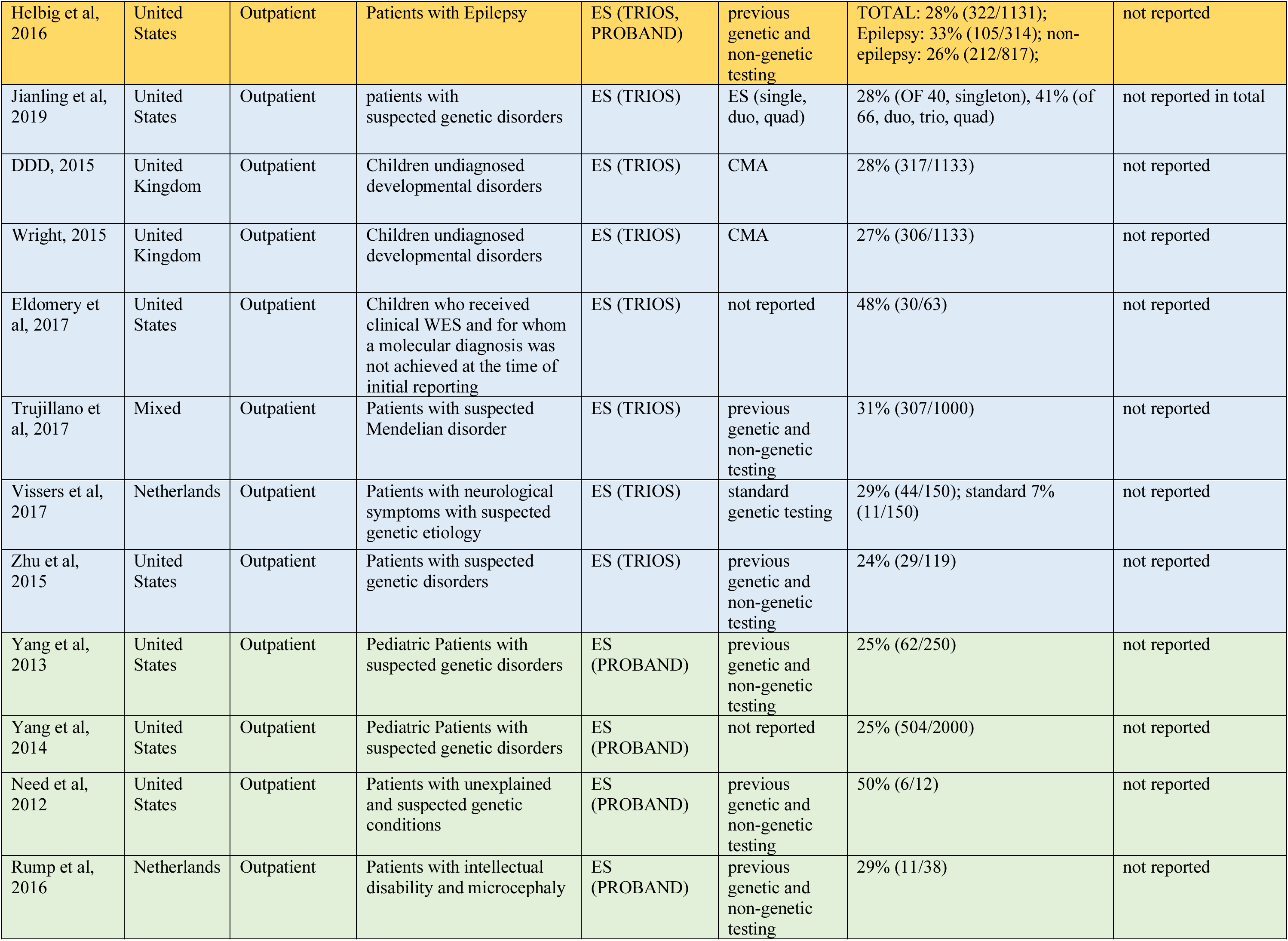
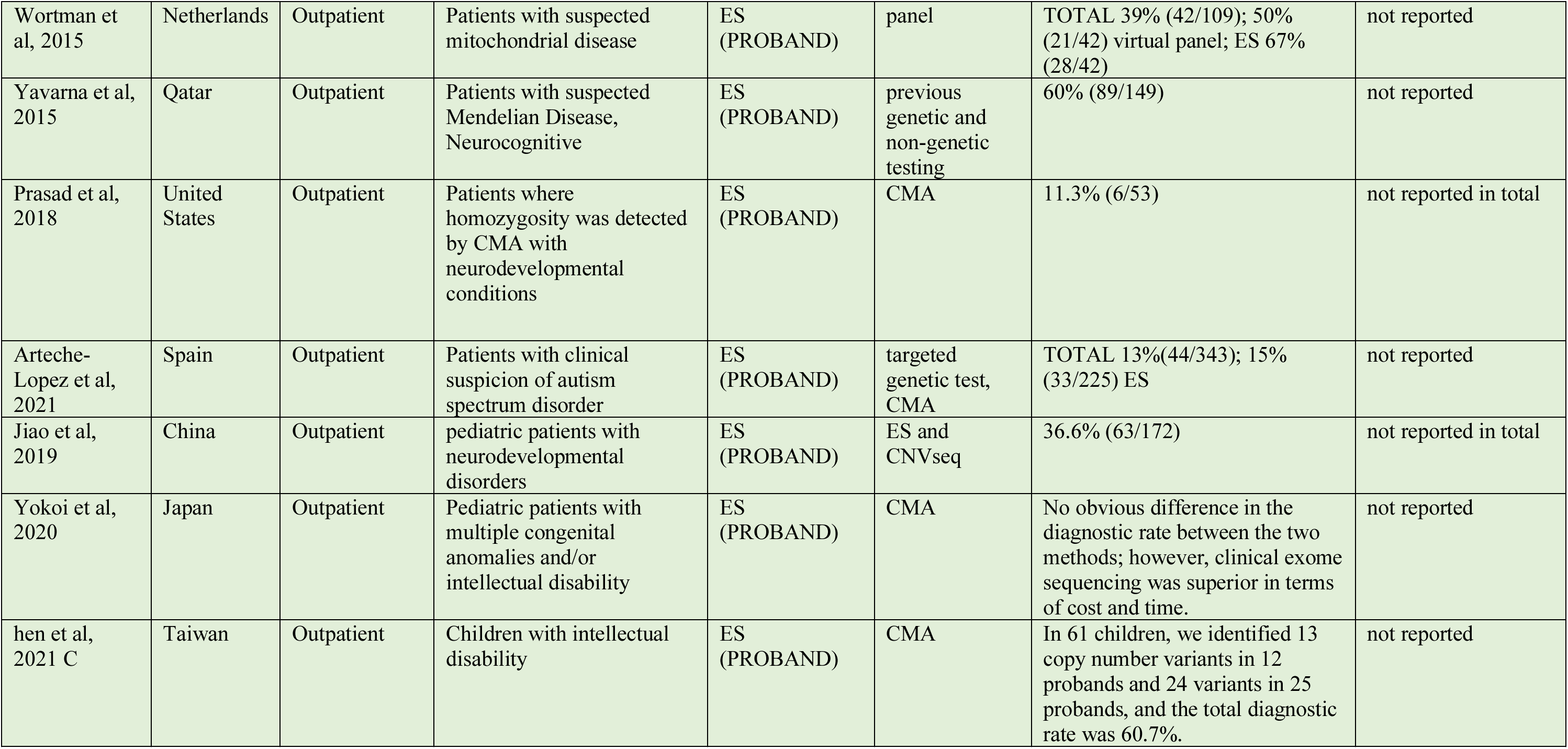
Studies that only reported Diagnostic Yield (n=28). Shading represents grouping by sequencing test type.

| Lastname, year | Site | Setting | Population | Sequencing Test | Comparator | Diagnostic Yield | Change in Clinical Management |
| --- | --- | --- | --- | --- | --- | --- | --- |
| Lionel et al, 2017 | Canada | Outpatient | Pediatric patients with phenotype suggestive of genetic disorder | GS (PROBAND, TRIO) | standard genetic testing | 41% (42/103), 24% (25/103) standard | not reported |
| Liu et al, 2019 | China | Outpatient | Patients with 16 distinct rare diseases | GS (PROBAND) | previous genetic and non-genetic testing | 79 individuals from 16 independent families were performed for whole-genome sequencing (WGS) in an effort to identify the causative mutations for 16 distinct rare diseases. disease-causing variants were identified in 10 of the 16 investigated diseases, yielding a diagnostic rate of 62.5%. | not reported |
| Alfares et al, 2018 | Saudi Arabia | Outpatient | Patients with negative array comparative genomic hybridization and negative or inconclusive WES results before WGS was performed | GS (PROBAND) | CMA, ES | 9% (10/108), decreased after reanalysis to 7% (7/108) | not reported |
| Bowling et al, 2017 | United States | Outpatient | Children with developmental delay or intellectual disability | ES, GS (TRIOS, DUO, PROBAND) | not reported | Total: 27% (100/371) (27); Trio: 29% (90/309), Duo: 19% (8/42), Proband: 15% (3/20) | not reported |
| Farwell et al, 2015 | United States | Outpatient | Patients suspected genetic diseases | ES (PROBAND, TRIO) | single and trio | 21% (14/68, singleton), 37% (82/220, trio) | not reported |
| Lee et al, 2014 | United States | Outpatient | Patients with suspected genetic conditions | ES (PROBAND, TRIO) | previous genetic and non-genetic testing | TOTAL 26%(213/814); Trio: 31% (127/410), Singleton 22% (74/338) | not reported |
| Charng et al, 2016 | SA | Outpatient | Patients from families with brain malformation and/or developmental delay/intellectual disability. | ES (PROBAND, TRIOS) | not reported | 55% (17/31) | not reported |
| Monies et al, 2017 | SA | Outpatient | Patients with suspected Mendelian phenotypes | ES (PROBAND, TRIO) | panels | TOTAL 43% (149/347) | not reported |
| Retterer et al, 2016 | United States | Outpatient | Patients with many different clinical indications suspected genetic disorders | ES (PROBAND, TRIO) | not reported | 21% (69/332, singleton), 31% (521/1686, trio) | not reported |
| Helbig et al, 2016 | United States | Outpatient | Patients with Epilepsy | ES (TRIOS, PROBAND) | previous genetic and non-genetic testing | TOTAL: 28% (322/1131); Epilepsy: 33% (105/314); non-epilepsy: 26% (212/817); | not reported |
| Jianling et al, 2019 | United States | Outpatient | patients with suspected genetic disorders | ES (TRIOS) | ES (single, duo, quad) | 28% (OF 40, singleton), 41% (of 66, duo, trio, quad) | not reported in total |
| DDD, 2015 | United Kingdom | Outpatient | Children undiagnosed developmental disorders | ES (TRIOS) | CMA | 28% (317/1133) | not reported |
| Wright, 2015 | United Kingdom | Outpatient | Children undiagnosed developmental disorders | ES (TRIOS) | CMA | 27% (306/1133) | not reported |
| Eldomery et al, 2017 | United States | Outpatient | Children who received clinical WES and for whom a molecular diagnosis was not achieved at the time of initial reporting | ES (TRIOS) | not reported | 48% (30/63) | not reported |
| Trujillano et al, 2017 | Mixed | Outpatient | Patients with suspected Mendelian disorder | ES (TRIOS) | previous genetic and non-genetic testing | 31% (307/1000) | not reported |
| Vissers et al, 2017 | Netherlands | Outpatient | Patients with neurological symptoms with suspected genetic etiology | ES (TRIOS) | standard genetic testing | 29% (44/150); standard 7% (11/150) | not reported |
| Zhu et al, 2015 | United States | Outpatient | Patients with suspected genetic disorders | ES (TRIOS) | previous genetic and non-genetic testing | 24% (29/119) | not reported |
| Yang et al, 2013 | United States | Outpatient | Pediatric Patients with suspected genetic disorders | ES (PROBAND) | previous genetic and non-genetic testing | 25% (62/250) | not reported |
| Yang et al, 2014 | United States | Outpatient | Pediatric Patients with suspected genetic disorders | ES (PROBAND) | not reported | 25% (504/2000) | not reported |
| Need et al, 2012 | United States | Outpatient | Patients with unexplained and suspected genetic conditions | ES (PROBAND) | previous genetic and non-genetic testing | 50% (6/12) | not reported |
| Rump et al, 2016 | Netherlands | Outpatient | Patients with intellectual disability and microcephaly | ES (PROBAND) | previous genetic and non-genetic testing | 29% (11/38) | not reported |
| Wortman et al, 2015 | Netherlands | Outpatient | Patients with suspected mitochondrial disease | ES (PROBAND) | panel | TOTAL 39% (42/109); 50% (21/42) virtual panel; ES 67% (28/42) | not reported |
| Yavarna et al, 2015 | Qatar | Outpatient | Patients with suspected Mendelian Disease, Neurocognitive | ES (PROBAND) | previous genetic and non-genetic testing | 60% (89/149) | not reported |
| Prasad et al, 2018 | United States | Outpatient | Patients where homozygosity was detected by CMA with neurodevelopmental conditions | ES (PROBAND) | CMA | 11.3% (6/53) | not reported in total |
| Arteche-Lopez et al, 2021 | Spain | Outpatient | Patients with clinical suspicion of autism spectrum disorder | ES (PROBAND) | targeted genetic test, CMA | TOTAL 13%(44/343); 15% (33/225) ES | not reported |
| Jiao et al, 2019 | China | Outpatient | pediatric patients with neurodevelopmental disorders | ES (PROBAND) | ES and CNVseq | 36.6% (63/172) | not reported in total |
| Yokoi et al, 2020 | Japan | Outpatient | Pediatric patients with multiple congenital anomalies and/or intellectual disability | ES (PROBAND) | CMA | No obvious difference in the diagnostic rate between the two methods; however, clinical exome sequencing was superior in terms of cost and time. | not reported |
| hen et al, 2021 C | Taiwan | Outpatient | Children with intellectual disability | ES (PROBAND) | CMA | In 61 children, we identified 13 copy number variants in 12 probands and 24 variants in 25 probands, and the total diagnostic rate was 60.7%. | not reported |

### Clinical Utility

From our search we identified 18 studies with NICU/PICU patients and 18 studies of outpatients that reported a change in management. In an effort to not duplicate the previous work of others (Stevens Smith, Clark), we report only the results from studies that are newer than these articles or that were not included in these articles. These include 13 studies in the NICU/PICU and five in the outpatient settings. The full set of all articles identified are in SUPPLEMENTAL TABLE 3 - ALL NICU-PICU Change in Management Studies (N=18), and SUPPLEMENTAL TABLE 4 - ALL Outpatient Change in Management Studies (N = 18).

**Table 4:**
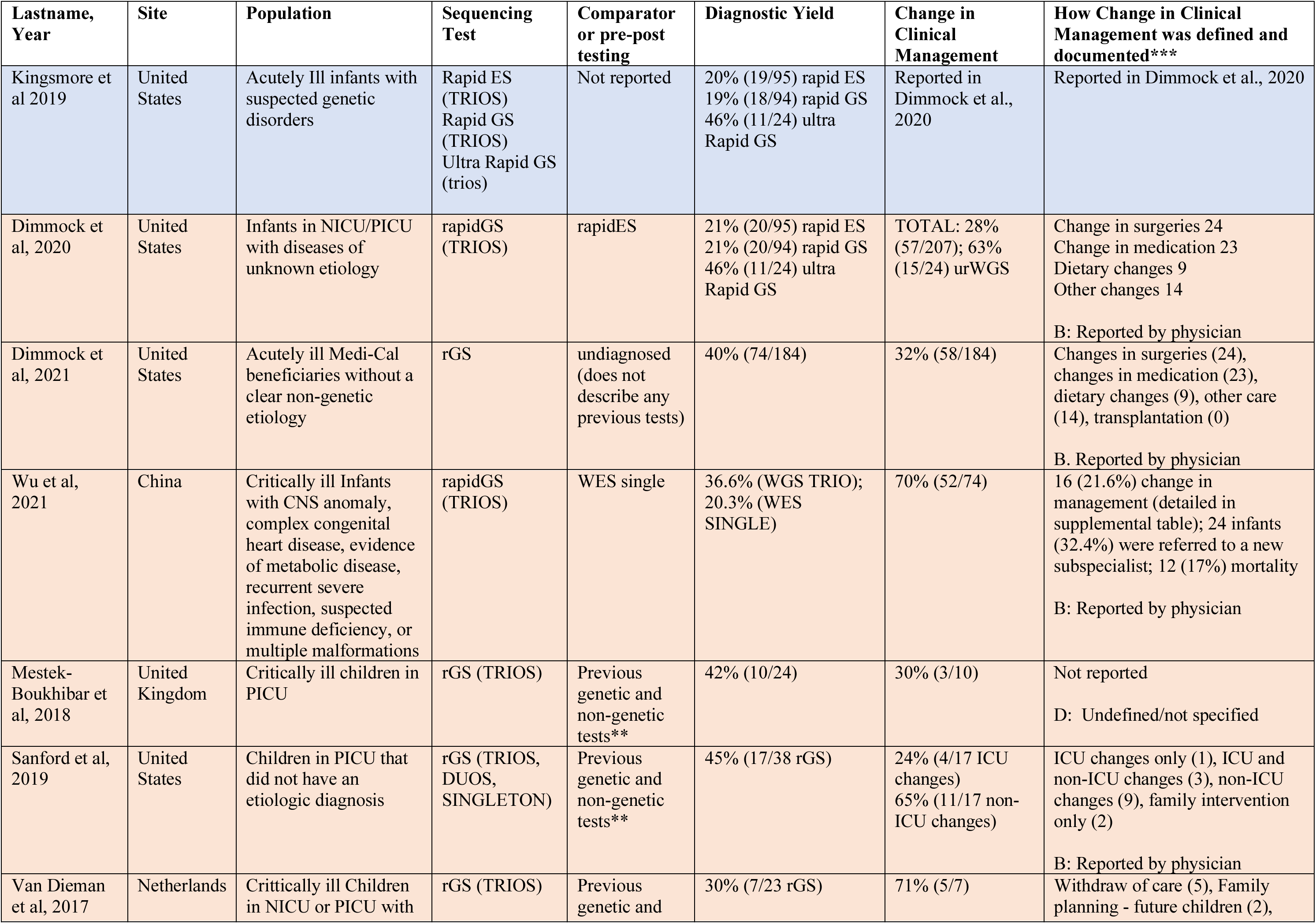
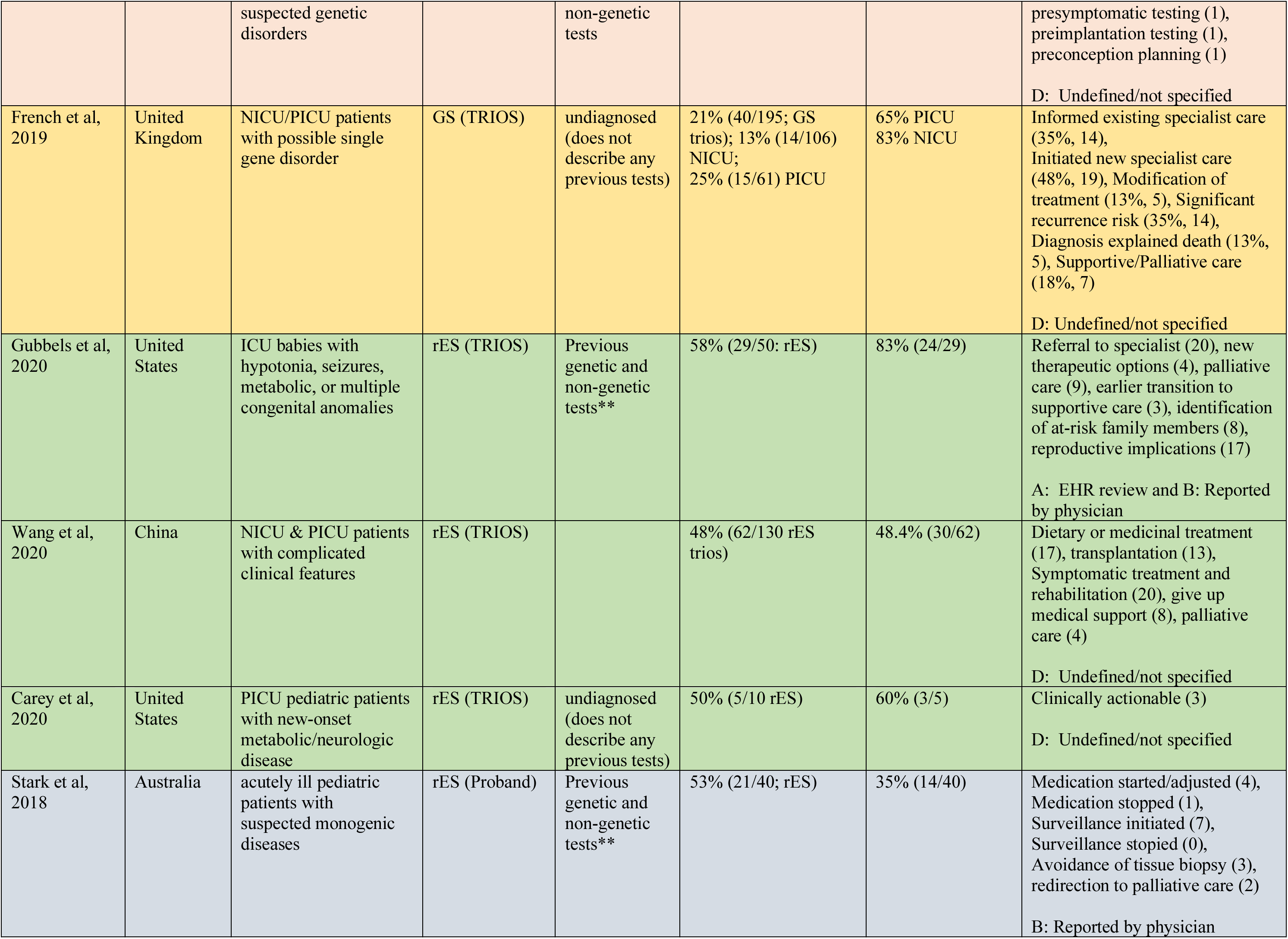
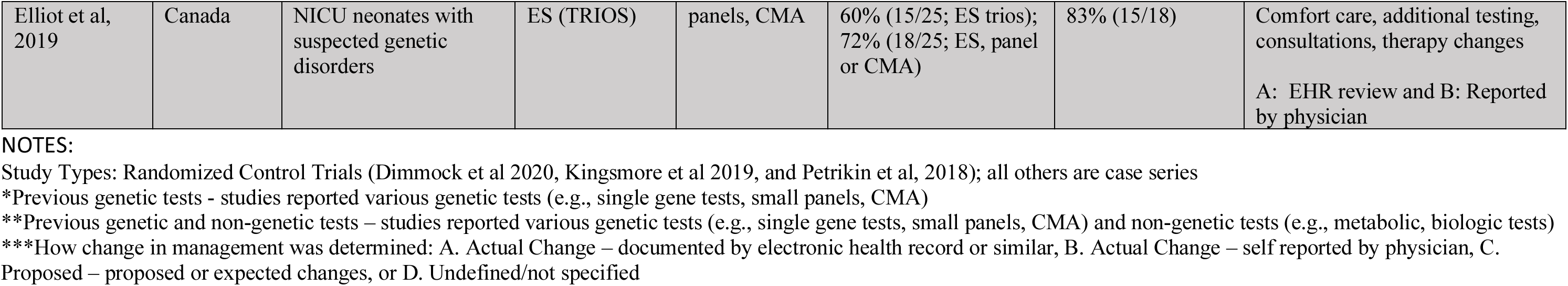
New studies of NICU/PICU (inpatient) setting reporting diagnostic yield and change in management (n = 13). Shading represents grouping by sequencing test type.

#### NICU/PICU setting

The majority (11/13) of studies reported the diagnostic yield and change in clinical management utilizing rapid GS or rapid ES, and one study each for GS and ES (See Table 4). One study reported on ultrarapid GS. All but four studies had less than 100 patients in total. The change in management was very heterogeneous ranging from 24% to 83% depending on the patients’ suspected disorder and test type (e.g., rapid GS, rapid ES, GS). We were unable to determine if any of the GS test types (ultra rapid GS or rapid GS or GS, range 24% - 83%) provided a greater change in clinical management rate than the ES test types (rapid ES or ES, range 35% - 83%).

#### Outpatient setting

All of the studies (N=3) reported the diagnostic yield and change in clinical management singleton ES (see table 5). The studies had less than 200 patients in total, with two of the studies with 100 or less patients. The change in management was very heterogeneous ranging from 6% to 61% depending on the patients’ suspected disorder.

**Table 5:**
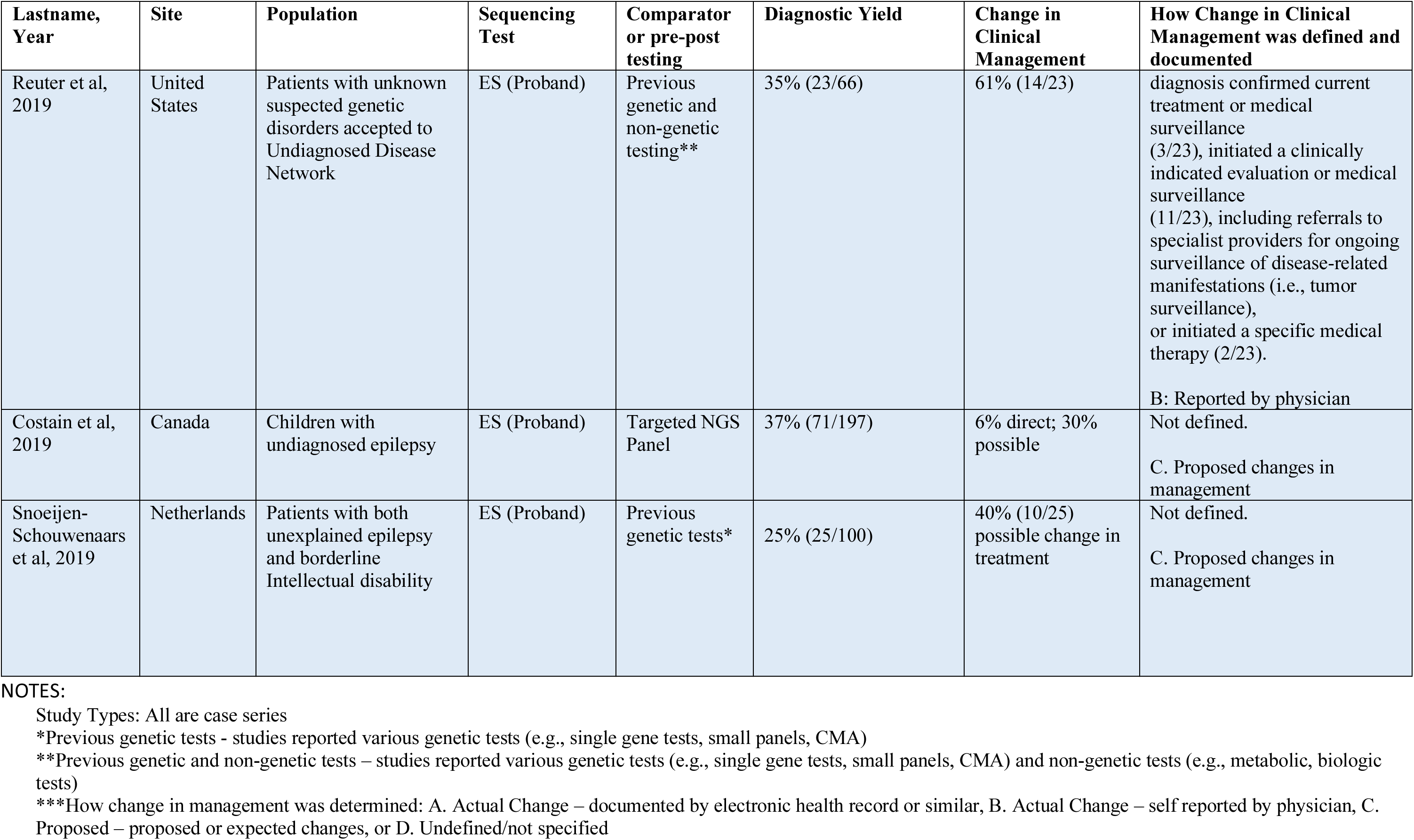
New studies of outpatient setting reporting diagnostic yield and/or change in management (n = 3). Shading represents grouping by sequencing test type.

#### Reporting of Change in Management

For both NICU/PICU and outpatient settings, the change in management was documented in the studies was heterogenous. How the actual change in management was described was extremely heterogenous with most studies using different descriptions for the types of change in management (see SUPPLEMENTAL TABLE 5: Detailed Methods and Type of Change in Management of Reporting (NICU/PICU and Outpatient) Genome and Exome Sequencing).

Most studies method of documenting the change in management using physician reporting (e.g., survey or interview) or electronic health records (See table 6). Few studies reported “proposed” or “intended” changes in management and many did not document how they determined the change in management.

**TABLE 6:**
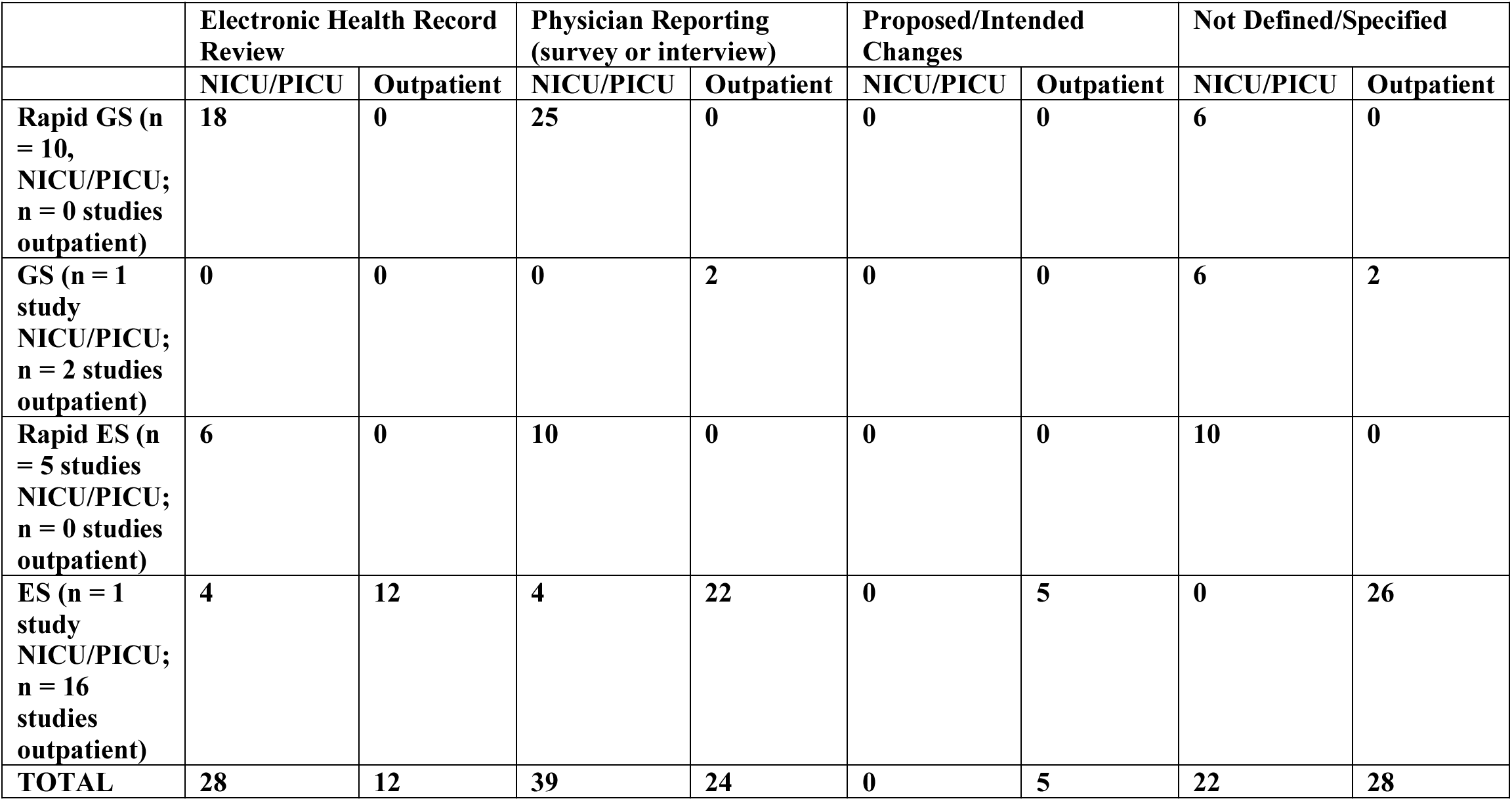
Total Change in Management Reporting by NICU/PICU and Outpatient Settings for Genome and Exome Sequencing.

|  | Electronic Health Record Review |  | Physician Reporting (survey or interview) |  | Proposed/Intended Changes |  | Not Defined/Specified |  |
| --- | --- | --- | --- | --- | --- | --- | --- | --- |
|  | NICU/PICU | Outpatient | NICU/PICU | Outpatient | NICU/PICU | Outpatient | NICU/PICU | Outpatient |
| Rapid GS (n = 10, NICU/PICU; n = 0 studies outpatient) | 18 | 0 | 25 | 0 | 0 | 0 | 6 | 0 |
| GS (n = 1 study NICU/PICU; n = 2 studies outpatient) | 0 | 0 | 0 | 2 | 0 | 0 | 6 | 2 |
| Rapid ES (n = 5 studies NICU/PICU; n = 0 studies outpatient) | 6 | 0 | 10 | 0 | 0 | 0 | 10 | 0 |
| ES (n = 1 study NICU/PICU; n = 16 studies outpatient) | 4 | 12 | 4 | 22 | 0 | 5 | 0 | 26 |
| **TOTAL** | **28** | **12** | **39** | **24** | **0** | **5** | **22** | **28** |

## Discussion

### Costs

We found limited data on the actual cost vs. price vs. list price for ES and GS. The cost or price varies based on the source. However, prices of tests alone underestimate the total cost of ES and GS but allow cross-study comparisons. The more rational measurement of comparison is done via cost-effectiveness review articles.

How costs are determined can make a difference in how health economic questions might be addressed. The use of a standard “cost” and cost-effectiveness analysis is a better way to consider whether it’s worth paying the higher price/cost of GS vs ES. The Lavelle article is the most current and comprehensive for our target population – pediatric patients suspected of genetic disorders.

Another consideration is reimbursement for GS or ES. Reimbursement for outpatient testing is typically billed to insurance as an individual item, whereas in the NICU/PICU setting all laboratory tests being lumped in with costs of a hospitalization. In the later, the hospital may end up being the responsible party for the majority of the cost of GS or ES.

### Comparative Effectiveness

We found limited evidence of a benefit of GS over ES, although there is a benefit over CMA and NGS Panels. The primary benefit appears not in the rate of change in management but in the timeliness of testing with rapid GS needed in the NICU/PICU vs. standard traditional GS or ES in the outpatient setting. Rapid GS or ES has a quick turn around of less than 14 days which is critical in the treatment of critically ill NICU/PICU patients. There is an expectation about the potential ability to take action on a given result (alteration of care in an acute setting versus longer term outpatient management of the diagnosed condition) to affect clinical outcomes.

Previous studies identified the ability of GS and ES to effect a change in management with varying results. The addition of the 13 NICU/PICU and 3 outpatient studies to the summary of evidence previously provided by Clark and Stevens Smith do not change the conclusions. There is no/limited evidence that GS is better than ES at identifying the ability to change clinical management.

Furthermore, there is a confounding factor of the ability of treating teams to perform adequate pre-test counseling and post-test results disclosure in a given inpatient/outpatient setting (taking into account that family members may or may not be available in each setting, and that inpatients can sometimes be discharged before test results come back raising questions about follow-up). Also, there is a difference in personal utility of genetic information for family members in acute versus chronic setting.

## Conclusion

There is limited evidence of GS over ES in the ability to effect change in management. Available evidence suggests that rapid over standard GS or ES is of greater benefit in the NICU/PICU setting vs. outpatient setting.

## Data Availability

All data produced in the present work are contained in the manuscript and supplemental materials

