## Supplemental for "Report – Cost and Clinical Utility of WES and WGS in pediatric patients with suspected genetic disease"

### Supplemental Materials

#### Supplemental Table 1: Search Strategies of Cost Studies

| | |
|---|---|
| Search String | (Genom* sequenc*[tiab] OR exom* sequenc*[tiab] OR WGS[tiab] OR WES[tiab]) AND ("Costs and Cost Analysis"[Mesh] OR Cost stud*[tiab] or cost differen*[tiab] or cost saving*[tiab] AND ((humans[Filter]) AND (english[Filter]))) |

#### Supplemental Table 2: Search Strategies of Comparative Effectiveness Studies

| | |
|---|---|
| Search String | Effectiveness Research"[Mesh] OR Clinical utility [tiab] AND ((humans[Filter]) AND (english[Filter]))) |

#### Supplemental Figure 1: Prisma Diagram of Comparative Effectiveness Studies

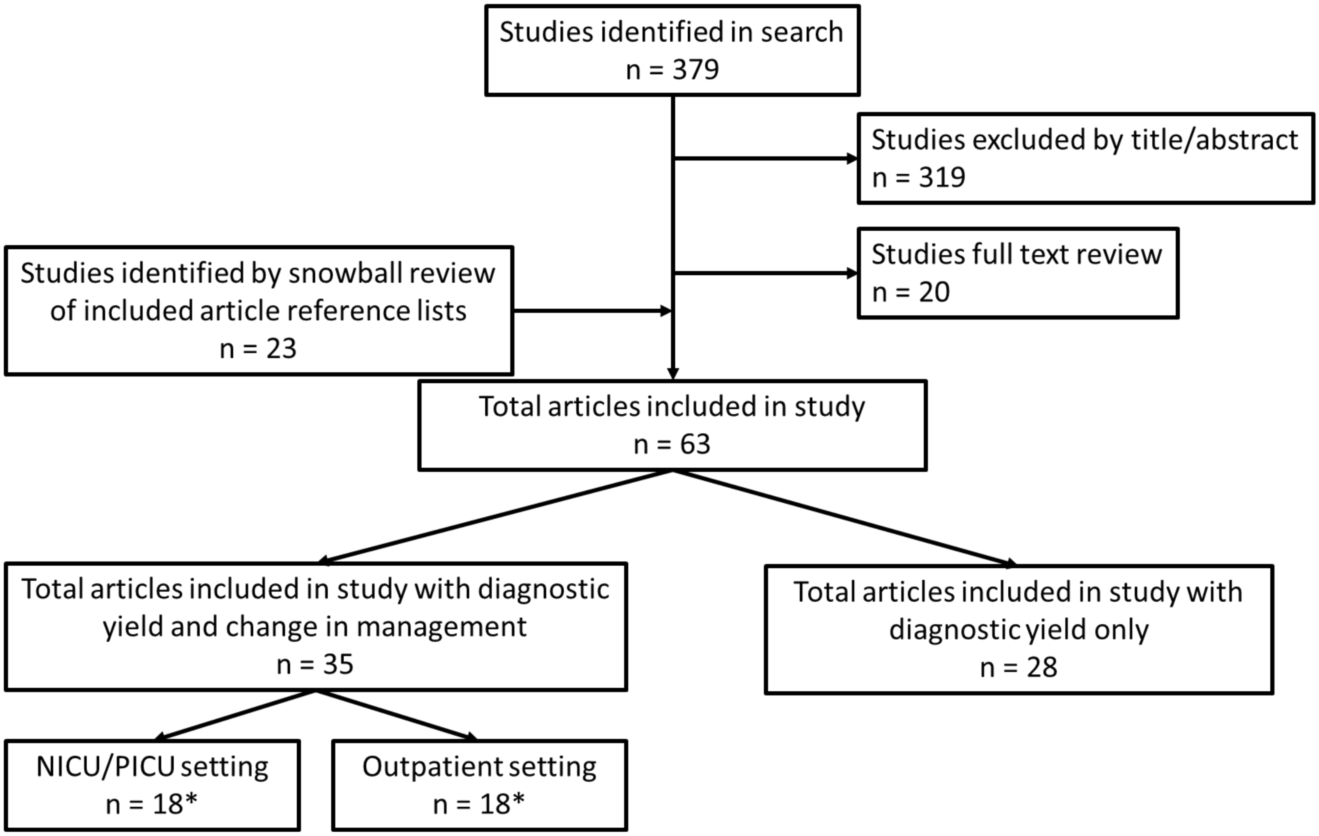

* One article (Soden) included patients in both settings

### Included Studies

#### Cost

#### Comparative Effectiveness

SUPPLEMENTAL TABLE 3 - **ALL** NICU-PICU Change in Management Studies (N=18)

**NOTE: Shading represents grouping by sequencing test type.**

| **Source of Article** | **Lastname, Year** | **Site** | **Population** | **Sequencing Test** | **Comparator or pre-post testing** | **Diagnostic Yield** | **Change in Clinical Management** | **How Change in Clinical Management was defined and documented*** ** |
|---|---|---|---|---|---|---|---|---|
| Dimmock | Kingsmore et al 2019 | United States | Acutely Ill infants with suspected genetic disorders | Rapid ES (TRIOS)<br>Rapid GS (TRIOS)<br>Ultra Rapid GS (trios) | Not reported | 20% (19/95) rapid ES<br>19% (18/94) rapid GS<br>46% (11/24) ultra Rapid GS | Same population as Dimmock et al, 2020 | Same population as Dimmock et al, 2020 |
| Clark | Farnaes et al. 2018 | United States | Acutely Ill infants with suspected genetic disorders | rapidGS (TRIOS) | Previous genetic tests* | 43% (18/42) | 31% (13/42) | Inotropic Support 7 (39%)<br>Respiratory Support 12 (67%)<br>Intubated 8 (44%)<br>Antimicrobial treatment 14 (78%)<br>>5 subspecialist consults 7(39%)<br>2 (11%) mortality<br><br>A: EHR review |
| Clark, Stevens Smith | Willig et al, 2015 | United States | Critically ill NICU and PICU infants withsuspected monogenetic disorder | rapidGS (TRIOS) | Previous genetic tests* | 57% (20/35); Standard 9% (3/32) | 65% (13/20) | Not defined<br><br>D: Undefined/not specified |
| Our Search | Dimmock et al, 2020 | United States | Infants in NICU/PICU with diseases of unknown etiology | rapidGS (TRIOS) | rapidES | 21% (20/95) rapid ES<br>21% (20/94) rapid GS<br>46% (11/24) ultra Rapid GS | TOTAL: 28% (57/207); 63% (15/24) urWGS | Change in surgeries 24<br>Change in medication 23<br>Dietary changes 9<br>Other changes 14<br><br>B: Reported by physician |
| Our Seach | Dimmock et al, 2021 | United States | Acutely ill Medi-Cal beneficiaries without a clear non-genetic etiology | rGS | undiagnosed (does not describe any previous tests) | 40% (74/184) | 32% (58/184) | Changes in surgeries (24), changes in medication (23), dietary changes (9), other care (14), transplantation (0) |

| | | | | | | | | |
|---|---|---|---|---|---|---|---|---|
| | | | | | | | | B. Reported by physician |
| Our Search | Wu et al, 2021 | China | Critically ill Infants with CNS anomaly, complex congenital heart disease, evidence of metabolic disease, recurrent severe infection, suspected immune deficiency, or multiple malformations | rapidGS (TRIOS) | WES single | 36.6% (WGS TRIO); 20.3% (WES SINGLE) | 70% (52/74) | 16 (21.6%) change in management (detailed in supplemental table); 24 infants (32.4%) were referred to a new subspecialist; 12 (17%) mortality<br><br>B: Reported by physician |
| Dimmock | Petrikin et al, 2018 | United States | Infants in NICU/PICU with illnesses of unknown etiology | rGS (TRIOS) | Previous genetic tests* | 31% (10/32 rWGS) 3% (1/33 SOC) | 31% (20/65) | Change in Management (14), genetic or reproductive counseling (13), Change other than counseling (8), change in subspecialty consult (5), medication change (1), procedure change (2), Diet Change (1). Imaging change (3), other (2)<br>A: EHR review and B: Reported by physician |
| Clark, Stevens Smith | Soden et al, 2014 (also reported as 2015) | United States | Children suspected of having a monogenetic disease | rapidGS (TRIOS) | undiagnosed (does not describe any previous tests) | 73% (11/15) rGS trio | 45% (22/49, trios) | New treatment (12), treatment discontinued(5), comorbidity evaluated(18), change in impression (12), other(11)<br><br>A: EHR review and B: Reported by physician |

| | | | | | | | | |
|---|---|---|---|---|---|---|---|---|
| Dimmock | Mestek-Boukhibar et al, 2018 | United Kingdom | Critically ill children in PICU | rGS (TRIOS) | Previous genetic and non-genetic tests** | 42% (10/24) | 30% (3/10) | Not reported<br><br>D: Undefined/not specified |
| Dimmock | Sanford et al, 2019 | United States | Children in PICU that did not have an etiologic diagnosis | rGS (TRIOS, DUOS, SINGLETON) | Previous genetic and non-genetic tests** | 45% (17/38 rGS) | 24% (4/17 ICU changes)<br>65% (11/17 non-ICU changes) | ICU changes only (1), ICU and non-ICU changes (3), non-ICU changes (9), family intervention only (2)<br><br>B: Reported by physician |
| Kingsmore | Van Dieman et al, 2017 | Netherlands | Crittically ill Children in NICU or PICU with suspected genetic disorders | rGS (TRIOS) | Previous genetic and non-genetic tests | 30% (7/23 rGS) | 71% (5/7) | Withdraw of care (5), Family planning - future children (2), presymptomatic testing (1), preimplantation testing (1), preconception planning (1)<br><br>D: Undefined/not specified |
| Dimmock | French et al, 2019 | United Kingdom | NICU/PICU patients with possible single gene disorder | GS (TRIOS) | undiagnosed (does not describe any previous tests) | 21% (40/195; GS trios); 13% (14/106) NICU;<br>25% (15/61) PICU | 65% PICU<br>83% NICU | Informed existing specialist care (35%, 14),<br>Initiated new specialist care (48%, 19), Modification of treatment (13%, 5), Significant recurrence risk (35%, 14),<br>Diagnosis explained death (13%, 5),<br>Supportive/Palliative care (18%, 7)<br><br>D: Undefined/not specified |
| Clark, Stevens Smith | Meng et al, 2017 | United States | Critically ill infants suspected to have genetic disorders | ES (Proband alone, and TRIOS) and | undiagnosed (does not describe any previous tests) | TOTAL 37% (102/278); 33% (58/178, singleton), | Standard ES (trio or proband): 52% (53/102) | **Standard ES (proband or trio)**<br>Redirection |

| | | | | | | | | |
|---|---|---|---|---|---|---|---|---|
| | | | | rapid ES (Trios) | | 44% (44/100, trio), Critical Trio 32/63 | Critical Trio: 71.9% (23/32) | of care (19/53), initiation of new subspecialist care (27/53), changes in medication or diet, (7/53), major procedures, such as a transplant (5/53)<br><br>**Critical Trio ES** Redirection of care (8/32), initiation of new subspecialist care (12/32), changes in medication or diet, (4/32), major procedures, such as a transplant (3/32)<br><br>D: Undefined/not specified |
| Dimmock | Gubbels et al, 2020 | United States | ICU babies with hypotonia, seizures, metabolic, or multiple congenital anomalies | rES (TRIOS) | Previous genetic and non-genetic tests** | 58% (29/50: rES) | 83% (24/29) | Referral to specialist (20), new therapeutic options (4), palliative care (9), earlier transition to supportive care (3), identification of at-risk family members (8), reproductive implications (17)<br><br>A: EHR review and B: Reported by physician |
| Dimmock | Wang et al, 2020 | China | NICU & PICU patients with complicated clinical features | rES (TRIOS) | | 48% (62/130 rES trios) | 48.4% (30/62) | Dietary or medicinal treatment (17), transplantation (13), Symptomatic treatment and rehabilitation (20), give up medical support (8), palliative care (4) |

| | | | | | | | | |
|---|---|---|---|---|---|---|---|---|
| | | | | | | | | D: Undefined/not specified |
| Dimmock | Carey et al, 2020 | United States | PICU pediatric patients with new-onset metabolic/neurologic disease | rES (TRIOS) | undiagnosed (does not describe any previous tests) | 50% (5/10 rES) | 60% (3/5) | Clinically actionable (3)<br><br>D: Undefined/not specified |
| Dimmock | Stark et al, 2018 | Australia | acutely ill pediatric patients with suspected monogenic diseases | rES (Proband) | Previous genetic and non-genetic tests** | 53% (21/40; rES) | 35% (14/40) | Medication started/adjusted (4), Medication stopped (1), Surveillance initiated (7), Surveillance stopied (0), Avoidance of tissue biopsy (3), redirection to palliative care (2)<br><br>B: Reported by physician |
| Our Search | Elliot et al, 2019 | Canada | NICU neonates with suspected genetic disorders | ES (TRIOS) | panels, CMA | 60% (15/25; ES trios); 72% (18/25; ES, panel or CMA) | 83% (15/18) | Comfort care, additional testing, consultations, therapy changes<br><br>A: EHR review and B: Reported by physician |

NOTES:

Study Types: Randomized Control Trials (Dimmock et al 2020, Kingsmore et al 2019, and Petrikin et al, 2018); all others are case series

*Previous genetic tests - studies reported various genetic tests (e.g., single gene tests, small panels, CMA)

**Previous genetic and non-genetic tests – studies reported various genetic tests (e.g., single gene tests, small panels, CMA) and non-genetic tests (e.g., metabolic, biologic tests)

***How change in management was determined: A. Actual Change – documented by electronic health record or similar, B. Actual Change – self reported by physician, C. Proposed – proposed or expected changes, or D. Undefined/not specified

SUPPLEMENTAL TABLE 4 - **ALL** Outpatient Change in Management Studies (N = 18)

**NOTE: Shading represents grouping by sequencing test type.**

| Source of Article | Lastname, Year | Site | Population | Sequencing Test | Comparator or pre-post testing | Diagnostic Yield | Change in Clinical Management | How Change in Clinical Management was defined and documented |
|---|---|---|---|---|---|---|---|---|
| Clark, Stevens Smith | Bick et al, 2017 | United States | Patients with suspected Mendelian disorder | GS (Proband) | not reported | After reanalysis: 36% (8/22)<br>Initial Analysis: 14% (3/22)<br>**[5 new cases for re-analysis]** | 75% (6/8) | Change in treatment and/or medical surveillance<br><br>B: Reported by physician |
| Clark, Stevens Smith | Stavropoulos et al, 2016 | Canada | Patients who met criteria for CMA by clinical geneticists and two or more structural malformations or unexplained developmental delay or intellectual disability | GS (Proband) | CMA + targeted sequencing, CMA alone | GS: 34% (34/100)<br>CMA + targeted panel: 13% (13/100)<br>CMA only: 8% (8/100) | 94% (32/34) | Category 1 (Disease-specific published management guidelines), Category 2 (Management based on case reports or known function of genes)<br><br>D: Undefined/not specified |
| Clark, Stevens Smith | Soden et al, 2014 | United States | Children suspected of having a monogenetic disease | ES (TRIOS) | not reported | ES Single: 45% (53/119)<br>ES Trio: 45% (45/100)<br>ES Trio (ambulatory): 40% (34/85)<br>GS after negative ES (1) | 45% (22/49, trios) | New treatment (12), treatment discontinued(5), comorbidity evaluated(18), change in impression (12), other(11)<br><br>A: EHR Review and B: Reported by physician |
| Clark, Stevens Smith | Valencia et al, 2015 | United States | Patients who had undiagnosed suspected genetic disease | ES (TRIOS) | Previous genetic and non-genetic testing** | 30% (12/40) | 100% (12/12) | Ending diagnostic odyssey, starting therapy or surveillance, |

| | | | | | | | | |
|---|---|---|---|---|---|---|---|---|
| | | | | | | | | informative genetic counseling<br><br>D: Undefined/not specified |
| Clark, Stevens Smith | Thevenon et al, 2016 | France | Patients with Intellectual Disorder and/or epileptic encephalopathy | ES (Proband) | Previous genetic and non-genetic testing** | Overall: 33% (14/43); Within Familial group: 67% (6/9) | 14% (2/14) | "personalization of care"<br><br>D: Undefined/not specified |
| Clark | Baldridge et al, 2017 | United States | Patients with suspected genetic disorders | ES (Proband) | not reported | 43% (67/155) | 12% (8/67) | Change or new treatment, surgery or other procedure, referral to specialist.<br><br>A: EHR review and B: Reported by physician |
| Clark, Stevens Smith | Iglesias et al, 2014 | United States | Patients with suspected genetic disorders (primarily birth defects and developmental delay) | ES (Proband) | Previous genetic and non-genetic testing** | 32% (37/115) | 65% (24/37) | Additional screening (8), altered management (14), therapy (2), familial mutation carriers (5), reproductive planning (6)<br><br>D: Undefined/not specified |
| Clark | Kuperberg et al, 2016 | Israel | Pediatric neurological patients suspected as having a monogenic disorder | ES (Proband) | Previous genetic and non-genetic testing** | 49% (28/57) | 18% (5/28) | Change in therapy (discontinuation or new), Dietary changes<br><br>A: EHR review |
| Clark, Stevens Smith | Srivastava et al, 2014 | Unites States | Pediatric patients with neurodevelopmental disorders | ES (Proband) | Previous genetic and non-genetic testing** | 41% (32/78) | 100% (32/32) | reproductive planning (n527), disease monitoring initiation (n54), |

| | | | | | | | | |
|---|---|---|---|---|---|---|---|---|
| | | | | | | | | investigation of systemic involvement of the disorder(s) (n56), alteration of presumed disease inheritance pattern (n57), changing of prognosis (n510), medication discontinuation (n55) or initiation (n52), and clinical trial education (n53).<br><br>D: Undefined/not specified |
| Clark, Stevens Smith, Our Search | Stark et al, 2016 | Australia | Pediatric patients with suspected monogenetic disorder | ES (Proband) | Previous genetic testing* | ES: 58% (46/80)<br>Standard Testing: 14% (11/80) | 33% (15/46) | Treatment (3), treatment discontinuation (1), treatment modifications (4), additional surveillance (9), discontinuation of surveillance (1).<br><br>B: Reported by physician |
| Clark, Stevens Smith | Tan et al, 2017 | Australia | Patients with suspected monogenic disorders | ES (Proband) | Previous genetic testing* | 52% (23/44) | 26% (6/23) | Not defined<br><br>A: EHR review |
| Clark, Stevens Smith | Tarailo-Graovac et al, 2016 | Canada | Patients with potential intellectual disability with metabolic phenotype of unknown cause | ES (Proband) | Previous genetic and non-genetic testing** | 68% (28/41) | 44% (18/41) | Preventative measures (4), therapy new or changes (15)<br><br>D: Undefined/not specified |

| | | | | | | | | |
|---|---|---|---|---|---|---|---|---|
| Stevens Smith | Nolan et al, 2016 | United States | Patients referred with suspected undiagnosed neurological disorders | ES (Proband) | Previous genetic and non-genetic testing** | 48% (24/50) | 33% (8/24) | family planning, medication selection, and systemic investigation<br><br>B: Reported by physician |
| Stevens Smith | Sawyer et al, 2015 | Canada | Patients suspected to have a genetic disorder | ES (Proband) | Previous genetic and non-genetic testing** | 29% (105/362) | 6% (6/105) | Therapy changes (3), Therapy initiation (3).<br><br>B: Reported by physician |
| Stevens Smith, Our Search | Tammimies et al, 2015 | Canada | Children with Autism Spectrum Disorder | ES (Proband) | CMA | ES: 8% (8/95)<br>CMA: 9% (24/258) | 6.2% (6/95) | additional baseline clinical investigations or ongoing screening are expected to improve outcome with respect to morbidity or mortality<br><br>C. Proposed changes in management |
| Our Search | Reuter et al, 2019 | United States | Patients with unknown suspected genetic disorders accepted to Undiagnosed Disease Network | ES (Proband) | Previous genetic and non-genetic testing** | 35% (23/66) | 61% (14/23) | diagnosis confirmed current treatment or medical surveillance (3/23), initiated a clinically indicated evaluation or medical surveillance (11/23), including referrals to specialist |

| | | | | | | | | |
|---|---|---|---|---|---|---|---|---|
| | | | | | | | | providers for ongoing surveillance of disease-related manifestations (i.e., tumor surveillance), or initiated a specific medical therapy (2/23).<br><br>B: Reported by physician |
| Our Search | Costain et al, 2019 | Canada | Children with undiagnosed epilepsy | ES (Proband) | Targeted NGS Panel | 37% (71/197) | 6% direct; 30% possible | Not defined.<br><br>C. Proposed changes in management |
| Our Search | Snoeijen-Schouwenaars et al, 2019 | Netherlands | Patients with both unexplained epilepsy and borderline Intellectual disability | ES (Proband) | Previous genetic tests* | 25% (25/100) | 40% (10/25) possible change in treatment | Not defined.<br><br>C. Proposed changes in management |

NOTES:

Study Types: All are case series

*Previous genetic tests - studies reported various genetic tests (e.g., single gene tests, small panels, CMA)

**Previous genetic and non-genetic tests – studies reported various genetic tests (e.g., single gene tests, small panels, CMA) and non-genetic tests (e.g., metabolic, biologic tests)

***How change in management was determined: A. Actual Change – documented by electronic health record or similar, B. Actual Change – self reported by physician, C. Proposed – proposed or expected changes, or D. Undefined/not specified

SUPPLEMENTAL TABLE 5: Detailed Methods and Type of Change in Management of Reporting (NICU/PICU and Outpatient) Genome and Exome Sequencing

| | Electronic Health Record Review | | Physician Reporting (survey or interview) | | Proposed/Intended Changes | | Not Defined/Specified | |
|---|---|---|---|---|---|---|---|---|
| | NICU/ PICU | Outpatient | NICU/ PICU | Outpatient | NICU/ PICU | Outpatient | NICU/ PICU | Outpatient |
| **Rapid GS (n = 10, NICU/PICU; n = 0 studies outpatient)** | | | | | | | | |
| Inotropic Support | 1 | | | | | | | |
| Respiratory support | 1 | | | | | | | |
| Intubated | 1 | | | | | | | |
| Antimicrobial treatment | 1 | | | | | | | |
| New treatment | 1 | | 1 | | | | | |
| Treatment discontinued | 1 | | 1 | | | | | |
| Subspecialist consults | 2 | | 2 | | | | | |
| Change in Surgery | | | 2 | | | | | |
| Transplant | | | 1 | | | | | |
| Change in medication | 1 | | 3 | | | | | |
| Diet Changes | 1 | | 2 | | | | | |
| Change in management | 1 | | 1 | | | | | |
| Procedure Change | 1 | | 1 | | | | | |
| Family intervention only (planning) | | | 1 | | | | 1 | |
| Presymptomatic testing | | | | | | | 1 | |
| Preimplantation testing | | | | | | | 1 | |
| Reproductive counseling | 1 | | 1 | | | | 1 | |
| Other counseling | 1 | | 1 | | | | | |
| Imaging Change | 1 | | 1 | | | | | |
| Comorbidity evaluated | 1 | | 1 | | | | | |
| ICU and non-ICU changes | | | 1 | | | | | |
| Non-ICU changes | | | 1 | | | | | |
| mortality | 1 | | 1 | | | | | |
| Withdrawal of care/Palliative care | | | | | | | 1 | |
| other | 1 | | 3 | | | | | |
| Not reported | | | | | | | 1 | |
| **GS (n = 1 study NICU/PICU; n = 2 studies outpatient)** | | | | | | | | |
| Change in treatment | | | | **1** | | | **1** | |

| | | | | | | | | |
|---|---|---|---|---|---|---|---|---|
| Informed existing specialist care | | | | | | | 1 | |
| New specialist care | | | | | | | 1 | |
| Change in Medical Surveillance | | | | **1** | | | | |
| Published management guidelines | | | | | | | | **1** |
| Management by published case reports | | | | | | | | **1** |
| Palliative care | | | | | | | **1** | |
| Mortality explained | | | | | | | **1** | |
| Risk recurrence | | | | | | | **1** | |
| **Rapid ES (n = 5 studies NICU/PICU; n = 0 studies outpatient)** | | | | | | | | |
| Redirection of care | | | | | | | **1** | |
| Clinically actionable | | | | | | | **1** | |
| Symptom treatment and rehabilitation | | | | | | | **1** | |
| Referral to specialist | **1** | | **1** | | | | | |
| New subspecialist care | | | | | | | **1** | |
| New therapeutic options | **1** | | **1** | | | | | |
| Changes in medication or diet | | | **1** | | | | **2** | |
| Major procedures (e.g., transplant) | | | | | | | **2** | |
| Surveillance initiated | | | **1** | | | | | |
| Surveillance stopped | | | **1** | | | | | |
| Avoidance of tissue biopsy | | | **1** | | | | | |
| Earlier transition to supportive care | **1** | | **1** | | | | | |
| Palliative care | **1** | | **1** | | | | **1** | |
| Give up medical support | | | | | | | **1** | |
| Identification of at-risk family members | **1** | | **1** | | | | | |
| Reproductive implications | **1** | | **1** | | | | | |
| **ES (n = 1 study NICU/PICU;n = 16 studies outpatient)** | | | | | | | | |
| Ending diagnostic Odyssey | | | | | | | | **1** |
| New Treatment | | **1** | | **1** | | | | **1** |

| | | | | | | | | |
|---|---|---|---|---|---|---|---|---|
| Treatment Discontinued | | **1** | | **1** | | | | |
| Comorbidity evaluated | | **1** | | **1** | | | | |
| Change in impression | | **1** | | **1** | | | | |
| Surveillance | | | | | | | | **1** |
| Genetic Counseling | | | | | | | | **1** |
| Other | | **1** | | **1** | | | | |
| “Personalization of Care” | | | | | | | | **1** |
| New Treatment | | **1** | | **2** | | | | **1** |
| Treatment Change | | **1** | | **3** | | | | |
| Change in Therapy (new or discontinuation) | **1** | **1** | **1** | **1** | | | | **2** |
| Treatment discontinuation | | | | **1** | | | | |
| Therapy | | | | | | | | **1** |
| Novel Therapy Options | | | | | | | | **1** |
| Medication Selection | | | | **1** | | | | |
| Change in medication or diet | | | | | | | | **1** |
| Dietary Changes | | **1** | | | | | | |
| Referrals to specialist for ongoing surveillance or specific medical therapy | | | | **1** | | | | |
| Referral to Specialist | **1** | **1** | **1** | **1** | | | | |
| New subspecialist care | | | | | | | | **1** |
| Diagnosis confirmed current treatment or medical surveillance | | | | **1** | | | | |
| Additional Surveillance | | | | **1** | | | | **1** |
| Discontinue Surveillance | | | | **1** | | | | |
| Started clinically indicated evaluation or surveillance | | | | **1** | | | | |
| Recommendations for clinical care | | | | | | | | **1** |
| Redirection of care/Comfort Care | **1** | | **1** | | | | | **1** |
| Change in prognosis | | | | | | | | **1** |
| Altered Management | | | | | | | | **1** |

| | | | | | | | | |
|---|---|---|---|---|---|---|---|---|
| Additional baseline clinical investigations | | | | | | **1** | | |
| Ongoing screening (expected to improve outcome w/ respect to morbidity/mortality) | | | | | | **1** | | |
| Additional Screening | | | | | | | | **1** |
| Additional Testing | **1** | | **1** | | | | | |
| Preventative Measures | | | | | | | | **1** |
| Familial Mutation Carriers | | | | | | | | **1** |
| Reproductive planning | | | | | | | | **2** |
| Family Planning | | | | **1** | | | | |
| Alteration of presumed disease inheritance pattern | | | | | | | | **1** |
| Major procedures (e.g. transplant) | | | | | | | | **1** |
| Investigation of systemic involvement of disorder | | | | **1** | | | | **1** |
| Clinical Trial education | | | | | | | | **1** |
| Other | | **1** | | **1** | | | | |
| Not Defined | | **1** | | | | **3** | | |
| **TOTAL** | **28** | **12** | **39** | **24** | **0** | **5** | **22** | **28** |
